# Autism spectrum disorders and brain volume link through a set of mTOR-related genes

**DOI:** 10.1101/2022.05.10.22274868

**Authors:** Martina Arenella, Nina R. Mota, Mariel W.A. Teunissen, Han G. Brunner, Janita Bralten

**Affiliations:** Department of Human Genetics, Radboud university medical center, Nijmegen, The Netherlands; Department of Forensic and Neurodevelopmental Science, Institute of Psychiatry, Psychology and Neuroscience, King’s College London, London, The United Kingdom; Donders Institute for Brain, Cognition and Behaviour, Radboud University, Nijmegen, The Netherlands; Department of Neurology, Maastricht university medical centre, Maastricht, The Netherlands; Department of Clinical Genetics, Maastricht University Medical centre, Maastricht, The Netherlands; GROW School of development and Oncology, and MHENS school of Neuroscience, Maastricht University, the Netherlands

**Keywords:** autism spectrum disorders, genetics, brain volume, mTOR, stratified genetic correlation

## Abstract

**Background:** Larger than average head and brain sizes are often observed in individuals with Autism Spectrum Disorders (ASDs). ASDs and brain volume are both highly heritable, with multiple genetic variants contributing. However, it is unclear whether ASDs and brain volume share any genetic mechanisms. Genes from the mammalian target of rapamycin (mTOR) pathway influence brain volume, and variants are found in rare genetic syndromes that include autism spectrum disorder features. Here we investigated whether variants in mTOR-related genes are also associated with ASDs and if they constitute a genetic link between large brains and ASDs.

**Methods:** We extended our analyses between large heads (macrocephaly) and rare *de novo* mTOR-related variants in an intellectual disability cohort (N=2,258). Subsequently using Fisher’s exact tests we investigated the co-occurrence of mTOR-related *de novo* variants and ASDs in the *denovo-db* database (N=23,098). We next selected common genetic variants within a set of 96 mTOR-related genes in genome-wide genetic association data of ASDs (N=46,350) to test gene-set association using MAGMA. Lastly, we tested genetic correlation between genome-wide genetic association data of ASDs (N=46,350) and intracranial volume (N=25,974) globally using LD-score regression as well as mTOR-specific by restricting the genetic correlation to the mTOR-related genes using GNOVA.

**Results:** Our results show that both macrocephaly and ASDs occur above chance-level in individuals carrying rare *de novo* variants in mTOR-related genes. We found a significant mTOR gene-set association with ASDs (*p*=0.0029) and an mTOR-stratified positive genetic correlation between ASDs and intracranial volume (*p*=0.027), despite the absence of a significant genome-wide correlation (*p*=0.81).

**Conclusions:** This work indicates that both rare and common variants in mTOR-related genes are associated with brain volume and ASDs and genetically correlate them in the expected direction. We demonstrate that genes involved in mTOR signalling are potential mediators of the relationship between having a large brain and having ASDs.

## Background

Autism spectrum disorders (ASDs) are a set of common neurodevelopmental conditions that are clinically characterized by atypical social communication and interaction, the presence of rigid, repetitive, and stereotyped behaviors, and atypical sensory processing (American Psychiatric Association, 2013). Heterogeneity at the phenotypic and etiological level in ASDs make it difficult to pinpoint the underlying biological mechanisms and to make steps towards better targeted (i.e., personalized) treatment options. One of phenotypic heterogeneities in ASDs is observed at the level of head size, with a larger than average head size being reported in some individuals with ASDs in the early years of life (Sacco et al., 2015). These individuals may present with macrocephaly and/or head circumference above the normal range (i.e., occipitofrontal circumference (OFC) > two standard deviations above the mean). The occurrence of large head circumference in ASDs is estimated to range between 12-15% and correlates with clinical severity(Fidler et al., 2000). Because head circumference is correlated with brain volume(Lindley et al., 1999), it may reflect a larger than average brain size. Previous studies indicate that individuals with ASDs have larger estimates of intracranial volume, total grey matter volume and cortical thickness in comparison with their matched controls(Van Rooij et al., 2018; Vonder Haar et al., 2017). The molecular mechanisms that may contribute to the observed variation in brain and head size in ASDs are not defined yet.

Previous studies suggest that genetic factors may play a role. Large heads are not only observed in individuals with ASDs but have been reported in ∼15% of their first-degree relatives(Butler et al., 2005). Both ASDs and brain volume have moderate-to-high heritability (h^2^) estimated around ∼70-90% (Tick et al., 2016) and ∼65%(Zhao et al., 2019) and their genetic architecture is complex and multifactorial – including both common and rare genetic variants. The question remains if these two traits share some degree of genetic liability. A previous study investigated the genetic correlation between ASDs and brain volume on a genome-wide scale and found no significant genetic correlation(Adams et al., 2016). However, it is possible that these two traits are connected through specific genetic variants and that such effects are diluted in global genetic correlation analyses (Werme et al., 2021)

Genetic studies of brain phenotypes report associations of multiple genes(Adams et al., 2016; Grasby et al., 2020), including genes controlling mTOR signalling (Licausi & Hartman, 2018), (Kim, 2015). Genes belonging to the mTOR signalling pathway are known to mediate general cell growth, proliferation, and survival(Yu & Cui, 2016) and are organized in two distinct branches, the RAS-MAPK sub-branch and the PI3K-AKT sub-branch(Kanehisa & Goto, 2000; Reijnders et al., 2017). During neurodevelopment, these mTOR-related genes also influence the proliferation of neuronal progenitor cells and are key contributors to the genesis of neurons and their reciprocal communication(Licausi & Hartman, 2018). The PI3K-AKT mTOR sub-branch controls the size of the soma and dendrite tree, whereas its interaction with the RAS-MAPK sub-branch allows the arborization of dendrites and the integration of synapses throughout the brain(Costa-Mattioli & Monteggia, 2013), (Kumar et al., 2005). Animal studies showed that dysregulation of mTOR signalling may drive excessive brain growth, that is consistent with a halt of neuronal apoptosis and consequent alterations in normal synaptic pruning(Huber et al., 2015). In a previous study *de novo* variants in a set of 101 mTOR-related genes were associated with the occurrence of macrocephaly in individuals with intellectual disability (ID). Moreover, common variants in this mTOR gene-set showed an association with intracranial volume (ICV) in the general population(Reijnders et al., 2017).

Some of the mTOR-related genes have also been implicated in ASDs(Onore et al., 2017). For example, animal studies showed that variants in the mTOR-related gene *PTEN* drive social impairment, a core feature of ASDs, in rodents (Tilot et al., 2016). Variants in *PTEN, MTOR* and *PIK3CA*, which are also mTOR-related genes, have been encountered in individuals with rare syndromes that comprise ASDs, and, in some instances, also macrocephaly(Klein et al., 2013), (Yeung et al., 2017). Besides rare syndromes, the mTOR-related genes have also been linked to idiopathic ASDs. The Simons Foundation Autism Research Initiative (SFARI) defined a list of genes based on both rare and idiopathic ASD studies and showed that many of these ASD-associated genes belong to the mTOR and the RAS-MAPK signalling gene-sets (Wen et al., 2016). Research on common genetic variants further indicated an over-representation of mTOR-related genes among ASD-associated genetic variants (The Autism Spectrum Disorders Working Group of The Psychiatric Genomics Consortium, 2017).

Taken together, there is evidence of the occurrence of increased head and brain size in ASDs and studies that investigated brain volume and ASDs separately both pointed to the potential contribution of mTOR-related genes. There is however no study that investigates whether genetic variants in mTOR-related genes represent a genetic link between ASDs and a tendency towards having a large head and brain. Therefore, the present study aims to investigate the specific role of genes from the mTOR gene-set in the relationship between brain volume and ASDs. We first explore locally collected data and publicly available data to investigate the incidence of macrocephaly and ASDs in individuals carrying rare *de-novo* variants in the set of 101 mTOR-related genes defined by Reijnders et al. 2017. Next, using common genetic variant data, we investigated whether the same mTOR gene-set, that previously showed an association with ICV, was also associated with ASD. Lastly, using common variant data for both ASD and ICV, we tested with a stratified genetic covariance analysis if the mTOR gene-set constitutes a specific genetic link between ASD and ICV variability and if this correlation was in the expected direction.

## Methods

The set of mTOR-related genes was selected based on the mTOR pathway representation in the Kyoto Encyclopaedia of Genes and Genomes(Kanehisa & Goto, 2000), with two sub-pathways: the PI3K-AKT-mTOR pathway and the RAS-MAPK-mTOR pathway (as defined by Reijnders et al., 2017). The list of mTOR-related genes was derived from three authoritative reviews of the mTOR regulators(Laplante & Sabatini, 2012; Shimobayashi & Hall, 2014; Sun & Hevner, 2014). Through literature search, protein complexes were mapped to single proteins and genes. The final list contains 101 mTOR-related genes: 96 genes map on autosomes, and 5 genes belong to the X-chromosome, with 60 genes belonging to the PI3K-AKT pathway and 76 genes to the RAS-MAPK pathway (*Table S1* for further details).

Following-up on previous work by Reijnders et al., 2017, we acquired additional exome data to reach a total of 2,258 patients with intellectual disability (ID). We aimed to confirm the enrichment of macrocephaly in patients carrying mTOR-related *de novo* variants. The participants had undergone diagnostic trio whole exome sequencing (WES) at the Radboud university medical center and the Maastricht University Medical Centre. Level of ID ranged between mild (ID 50-70) and severe (IQ<30). Patients were classified as macrocephalic (OFC > 2.5 SD), normocephalic (2.5 SD < OFC < 2.5 SD) or microcephalic (OFC < 2.5 SD). Diagnostic WES was approved by the local medical ethics committee. Written informed consent was obtained from all the families. Patient-parent trio exomes were sequenced, using DNA isolated from blood at the Bejing Genomics Institute in Copenhagen. Exome capture was performed using Agilent SureSelect v4 and v5 and samples were sequenced on an Illumina HiSeq 4000 platform with paired-end reads to a median target coverage of 112x. Sequence reads were aligned to the hg19 reference genome using BWA (v0.7.12). Variants were subsequently called by the GATK haplotypecaller (v3.4-46). For further details, see *Supplementary Information*. We performed a Fisher’s exact test in R to establish if the occurrence of macrocephaly was significantly higher in mTOR-related variant carriers compared to chance, setting the significance threshold at 0.05. Under- or over-enrichment of macrocephaly among mTOR-related variant carriers was tested based on the cumulative distribution function of the hypergeometric distribution using the webtool of the Graeber Lab, https://systems.crump.ucla.edu/hypergeometric/. Similarly, we tested specific enrichment for *de novo* variants in genes of the RAS-MAPK and PIK3-AKT gene sub-branches separately.

To understand whether *de novo* variants in the mTOR gene-set have been reported in ASDs, we exploited an online database of *de novo* variants, the *de-novo-db* database (http://denovo-db.gs.washington.edu/denovo-db/). This database includes 40 independent studies on psychiatric and neural phenotypes providing a list of genome-wide *de novo* variants detected in 23,098 individuals. All participants in each study provided written informed consents and agreed with the use of their data for research purposes. We filtered the *de-novo-db* data on participants diagnosed with “autism” (the definition used in *de-novo-db*) as primary phenotype and controls. We used the publicly available data that did not include data derived from the Simon Complex Sample Collection. For further details about diagnostic criteria and cohort descriptives in the *de-novo-db* see (Turner et al., 2017) and http://denovo-db.gs.washington.edu/denovo-db/. We counted *de novo* variants in the mTOR gene-set and in genes outside the set in both individuals with autism and in controls. As background we used genes throughout the genome and subsequently genes expressed in the brain (based on annotations from eMAGMA(Gerring et al., 2021). We built a 2x2 contingency table (*Table S2*) and performed a Fisher’s exact test to assess the (in)dependence between mTOR-related *de novo* variants and autism. Analyses were performed in R and the significance threshold was set at 0.05. Under- or over-enrichment of autism among carriers of variants in mTOR-related genes was tested based on the cumulative distribution function of the hypergeometric distribution using the webtool from the Graeber Lab, https://systems.crump.ucla.edu/hypergeometric/. As sensitivity analyses, we tested the enrichment of variants in genes from the RAS-MAPK sub-branch and the PIK3-AKT subbranch respectively in individuals with autism and in controls.

Next, we investigated if common genetic variants within the mTOR gene-set were linked to ASDs using the summary statistics resulting from the ASD genome-wide association study (GWAS) meta-analysis conducted by the Psychiatric Genomic Consortium (PGC) and the Lundbeck Foundation Initiative for Integrative Psychiatric Research (iPSYCH) that included 18,381 ASD cases and 27,969 controls(Grove et al., 2019; see *Supplementary information*). All the individuals included in the PGC and iPSYCH cohort samples provided written informed consent. We tested the full set of 96 autosomal mTOR-related genes, and post-hoc looked at the PI3K-AKT and the RAS-MAPK gene-sets. Gene-set analyses were performed using the Multi Maker Annotation for Genomic Annotation (MAGMA) software package (version 1.09)(de Leeuw et al., 2015) using the SNP-wise mean model that combines *p*-values of the gene-related SNPs, controlling for linkage disequilibrium (LD). In addition, we performed conditional analyses, where we conditioned on brain expressed genes considering that these genes are enriched for common variants associated with brain-related traits, such as ASDs and ICV(Brien et al., 2018). Gene-set results were considered significant if they reached the Bonferroni-corrected *p*-value threshold of *p* < 0.025 (i.e., corrected by the number of tests).

To investigate the genetic link between ASDs and ICV we analysed the summary statistics from the ASD-GWAS meta-analysis (18,381 ASD cases and 27,969 controls)(Grove et al., 2019) with the summary statistics resulting from the ICV-GWAS meta-analysis conducted by the Enhancing Neuro-Imaging Genetics Through Meta-Analysis (ENIGMA) and the Cohorts for Heart and Aging Research in Genomic Epidemiology (CHARGE) Consortia that included 25,974 individuals(Adams et al., 2016) and see *Supplementary Information*. All the individuals included in the ENIGMA and CHARGE cohort samples provided written informed consent. We estimated the genetic correlation between the summary statistics of the PGC-ASD GWAS and the ENIGMA-ICV GWAS using LD bivariate score (LDSC) regression as implemented in the LDSCORE regression software(Bulik-Sullivan et al., 2015). Analyses used pre-computed LD scores based on the 1000 Genome Project reference panel that well-suits European-centred GWASs (available at https://github.com/bulik/ldsc). LDSC-based analysis consisted of two steps: 1) converting summary statistics data to the LDSC format (data filtering and merging to the reference panel as shown in https://github.com/bulik/ldsc/wiki/Heritability-and-Genetic-Correlation), 2) estimating genetic correlation. The regression model did not include any regression intercept given the absence of sample overlap between the investigated datasets. A block jack-knife procedure over SNP data was used to estimate standard errors and calculate corresponding *p*-values. Results were considered significant at *p*<0.05.

To estimate whether ASDs and ICV were genetically correlated through mTOR-related genes and if so in which direction, we used the GeNetic cOVariance Analyzer (GNOVA) method (https://github.com/xtonyjiang/GNOVA)(Lu et al., 2017). The GNOVA approach allows to stratify the genetic covariance between two traits of interest by pre-defined functional annotation, in our case the mTOR gene-set. In addition, it provides stratified correlation results by normalising the covariance estimates. We created annotations for the full mTOR gene-set using the make_annot.py script available at (https://github.com/bulik/ldsc/). Then, using the PGC-ASD and ENIGMA-ICV summary statistics as input we performed the stratified genetic correlation analysis. Subsequently, we used the annotation for the two mTOR sub-branches and respectively tested them in secondary GNOVA-based ASD-ICV correlation analyses.

## Results

In the local ID cohort, we identified 39 *de novo* variants in mTOR-related genes. Macrocephaly was observed in 33% of the patients with mTOR-related variants, while it was reported in 6% of the patients with *de novo* variants in genes outside the mTOR-related gene-set (*Table 1*).

**Table 1.** Intelectual disability cohort descriptives

| **Cohort** | **N** | **Age**<br>**(mean)** | **Sex** | **Macrocephalic**<br>**(OFC > +**<br>**2SD)** | **Normocephalic**<br>**(-2SD < OFC <**<br>**+ 2SD)** | **Microcephalic**<br>**(OFC < -2SD)** |
| --- | --- | --- | --- | --- | --- | --- |
| Total | 2258 | 9 | 43,8%<br>female | 163 | 1,707 | 388 |
| Patients with *de novo* variants | 2144 | 10,3 | 48% female | 142 | 1,712 | 290 |
| Patients with *de novo* mTOR-related variants | 39 | 12,4 | 52% female | 13 | 19 | 7 |

OFC = occipitofrontal cortex, N=sample size, OFC >+ 2SD = occipitofrontal cortex bigger than 2 times the standard deviation from the mean, -2SD < OFC < +2SD = occipitofrontal cortex between 2 times the standard deviation bigger and smaller than the mean, OFC < -2SD = occipitofrontal cortex smaller than two times the standard deviation away from the mean

The Fisher’s exact test indicated that macrocephaly was significantly more frequent in carriers of *de novo* variants in mTOR-related genes than individuals carrying variants in genes outside the mTOR gene-set (OR= 8.9; CI: 3.9-19.61; *p*= 1.7e-7). The hypergeometric test also indicated an over-enrichment of macrocephaly in carriers of *de novo* mTOR-related variants (5.03-fold change, p-value = 5e-07). We did not observe differences in the frequency of *de novo* variants between the RAS-MAPK and PI3k-AKT gene sub-branches.

We observed microcephaly in 17% of the patients carrying *de novo* variants in mTOR-related genes. However, the frequency of microcephaly did not statistically differ between carriers of variants in mTOR-related genes and carriers of variants in genes not included in the mTOR gene-set (OR= 2.2; CI = 0.77-5.53; *p*=0.08, see *Table S2*).

Exploring the *de-novo-db* database, we counted a total of 34,389 *de novo* variants (7,623 in brain-expressed genes) reported across individuals with autism and controls. Of these variants, 135 variants (32 for brain-expressed genes) occurred in genes belonging to the mTOR gene-set and 97% (132/135) were reported in individuals with autism. Fisher’s exact test indicated a significantly higher incidence of *de novo* variants in the genes of the mTOR gene-set in individuals with autism than in controls (OR=2.06 CI: 1.90-2.24; *p*=2.2e-16; positive enrichment fold 1.06; hypergeometric p-value=0.01). Analyses restricted to the brain expressed genes showed a positive but non-significant enrichment of mTOR-related *de novo* variants in autism as compared to variants in other brain expressed genes (enrichment fold 1.02; p> 0.5) (*Table S4*). Sensitivity analyses on the two sub-branches indicated a significant enrichment of variants in genes of the RAS-MAPK sub-branch in autism (enrichment fold = 1.06, p= 0.01), but not for variants in the PIK3-AKT sub-branch genes (enrichment fold= 1.02; p = 0.06). There was no significant enrichment of variants in either of the two sub-branches in autism when restricting our analyses to genes expressed in the brain (*Table S4*). For further details about the *de novo* variants in mTOR-related genes from *de-novo-db* see *Table S*3.

MAGMA-based gene-set analysis showed a significant association of the full set of 96 autosomal mTOR-related genes with both ASDs (beta(SD)=0.23(0.08); *p*= 0.0033) and ICV (beta(SD)=0.17(0.08); *p*= 0.021). Conditioning analyses on brain expressed genes confirmed these associations (ASDs: beta=0.22; *p*=0.003, ICV: beta=0.17; *p*=0.02). Post-hoc analyses on the two mTOR-related sub-branches indicated a significant association of ASDs with the RAS-MAPK pathway (beta=0.28; *p*= 0.002; conditioned on brain-expressed *p*= 0.001) and of ICV with the PI3K-AKT pathway (beta=0.29; *p*=0.00425; conditioned on brain-expressed *p=*0.00434)(*Table 2*).

**Table 2.** MAGMA-based gene-set analyses in autism spectrum disorders and intracranial volume

| Trait | Gene-set | Beta | SD | P | Cond-P |
| --- | --- | --- | --- | --- | --- |
| ASDs | Full mTOR | 0.23 | 0.08 | 0.0029* | 0.0033* |
|  | PI3K-AKT | 0.17 | 0.11 | 0.059 | 0.79 |
|  | RAS-MAPK | 0.28 | 0.09 | 0.0014* | 0.0016* |
| ICV | Full mTOR | 0.17 | 0.08 | 0.021* | 0.021* |
|  | PI3K-AKT | 0.29 | 0.11 | 0.0042* | 0.0044* |
|  | RAS-MAPK | 0.14 | 0.09 | 0.07 | 0.07 |

ASDs=autism spectrum disorders, ICV=intracranial volume, SD=standard deviation, P=p-value, Cond-p=conditional p-value \*Bonferonni=corrected significant p-values

The LDSC-based analysis indicates no significant genome-wide genetic correlation between ASDs and ICV (ρg= 0.040, *p*= 0.81).

The GNOVA-based analysis of genetic correlation stratified by the mTOR-related gene-set revealed a significant positive genetic correlation between ASDs and ICV (rg=0.001; *p*=0.027). Subsequent post-hoc analyses revealed that the genetics of ASDs and ICV were significantly positively correlated when stratified by the RAS-MAPK pathway (rg=0.001; *p*=0.002), but not when stratified by the PI3K-AKT pathway (rg=0.0004; *p*=0.10) (*Table 3*).

**Table 3.** Genetic correlations between autism spectrum disorders and intracranial volume

| Genetic annotation | rg | SD | P |
| --- | --- | --- | --- |
| Full genome | 0.04 | 0.05 | 0.81 |
| mTOR gene-set | 0.001 | 0.0004 | 0.027* |
| RAS-MAPK sub-branch | 0.0012 | 0.0003 | 0.002* |
| PI3K-AKT sub-branch | 0.00040 | 0.00024 | 0.105 |

Rg=genetic correlation, SD=standard deviation p=p-value \*Bonferroni-corrected significant p-values

## Discussion

By relying on multiple data sources and a combination of analytical tools, our results support the hypothesis that genes involved in mTOR signalling constitute a genetic link between ASDs and brain volume. The observed mTOR-gene-set-based correlation between ASDs and brain volume occurs in the expected positive direction in line with the phenotypic association between large brain and head size and ASDs.

We extended and confirmed the over-representation of rare mTOR-related *de novo* variants in individuals with macrocephaly in line with previous work (Reijnders et al., 2017)using a subset of our local ID cohort and we found that *de novo* variants in mTOR-related genes are over-represented in individuals with autism. Additionally, we show that common variants in the mTOR gene-set are, next to being associated to ICV(Reijnders et al., 2017), also associated with ASDs. Lastly, we demonstrated that common variants in the mTOR gene-set mediate a positive genetic correlation between ASDs and ICV, while no ASDs-ICV correlation was detected at a genome-wide level (*Figure 1*).

**Figure 1.**
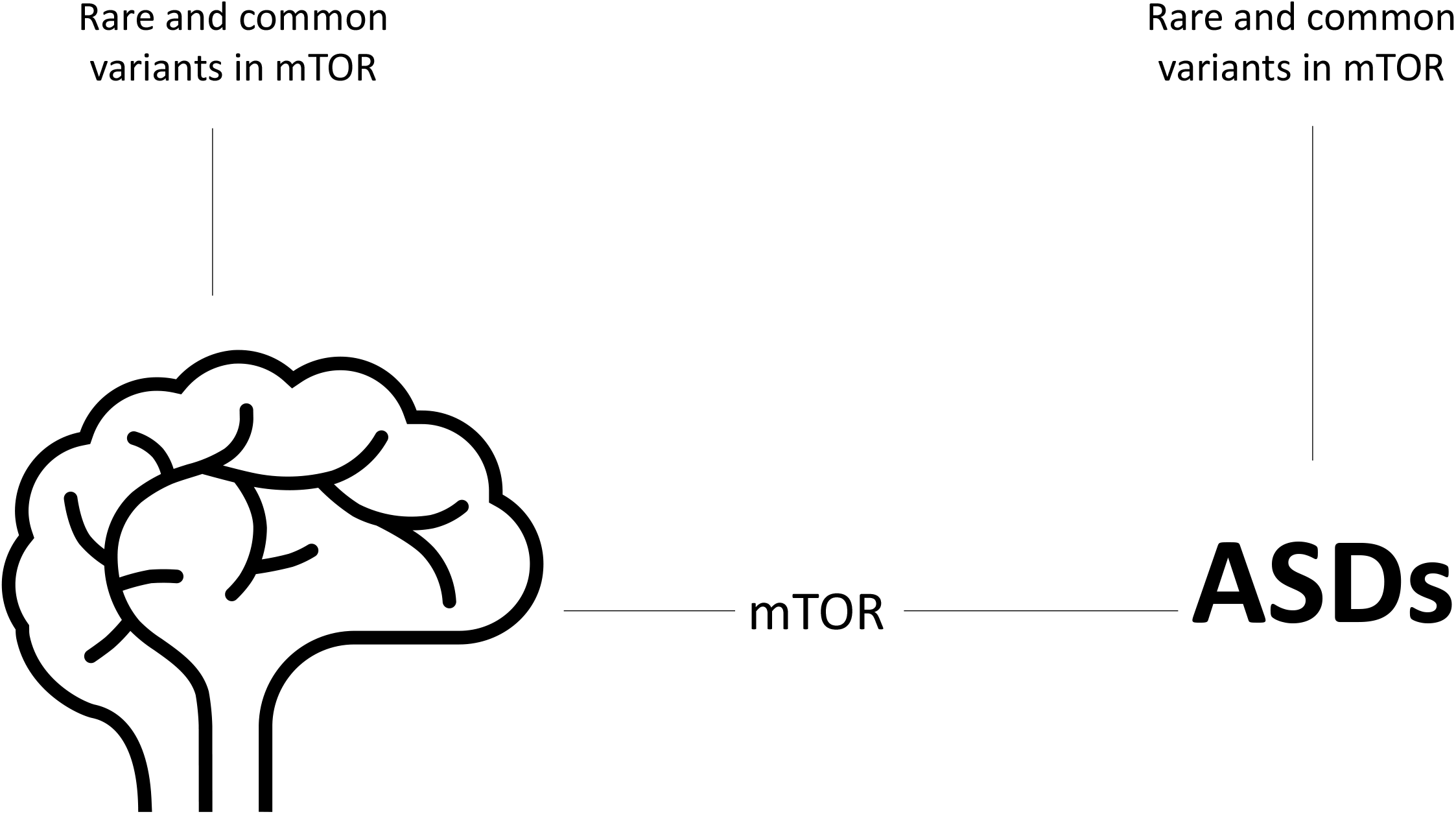
A phenotypic correlation between head and brain size and autism spectrum disorders (ASDs) is known, however no genome-wide genetic correlation was found between ASDs and intracranial volume (ICV). In this study we show that both rare and common variants in the mTOR gene-set link to brain volume and ASDs, and that stratifying the genetic correlation to variants within the mTOR gene-set leads to a significant positive genetic correlation between ASDs and ICV.

There is evidence for a phenotypic correlation between ASDs and large brains(Sacco et al., 2015), yet previous studies did not find significant genetic correlations between ASDs and brain volume(Grove et al., 2019). Our data support a more specific, positive genetic correlation through mTOR-related genes between ASDs and ICV, indicating an overlap between alleles that increase the odds of having ASDs and having a large brain. This provides a potential mechanistic underpinning of the clinical association of large brains in individuals with ASDs. These results highlight that stratification of genetic correlations can reveal that complex traits are linked specifically through functionally defined portions of the genome (Lu et al., 2017) and that this signal can be diluted when we look at the genetic correlation of the complete genome. Genetic stratification is particularly informative for those scenarios where genetic correlation between loci is confined to a specific genomic region, or when genetic factors show opposite effects at different loci that would cancel each other out in a standard, global-scale genetic correlation analyses(Werme et al., 2021). Dissecting the genetic relationship by function has the potential to delineate shared biological mechanisms between complex traits, such as brain volume and ASDs.

Our research focused on a specific set of mTOR-related genes, involving both RAS-MAPK and PI3K-AKT signalling, which have been studied separately in previous studies on ASDs and brain volume. The ENIGMA consortium, relying on the same ICV-GWAS data included in our study, showed that the top ICV-associated genetic variants were enriched for genes annotated to PI3K-AKT signalling(Adams et al., 2016). In line with those findings, we show here that the complete PI3K-AKT gene-set is associated with ICV. On the other hand, bioinformatic analysis of the ASDs risk genes from the SFARI database revealed an overrepresentation of genes within the RAS-MAPK mTOR sub-branch (Wen et al., 2016). Here we were able to expand on those studies by showing the positive genetic link between ASDs and ICV specifically for the RAS-MAPK pathway, and not for the PI3K-AKT pathway. These results were confirmed by our findings of a significant enrichment of *de novo* variants in the RAS-MAPK pathway in ASDs. Considering the functional role of these pathways, it is possible that the co-occurrence of ASDs and large ICV is explained by RAS-MAPK-mediated increased dendritic growth and spine formation, key to synaptic communication and regulated by the RAS-MAPK pathway (Kumar et al., 2005).

Our findings inform the need for further research on the mTOR signalling pathway as a potential target for treatment in ASDs. Previous studies already investigated mTOR inhibitors (e.g., rapamycin) in ASDs based on the evidence of up-regulation of the mTOR pathway in monogenic syndromes that include ASDs as well as in animal models(Schneider et al., 2017). Specifically, mTOR-related inhibitors appear effective in the rescue of neuronal and behavioural alterations across a range of ASD-related syndromes. Recently, Ganesan et al. (2019), reviewed the possibility to translate the use of mTOR inhibitors to the treatment of idiopathic ASDs, and highlight the downstream regulators of the mTOR signalling as valuable molecular targets(Ganesan et al., 2019). Our data support research on these mTOR inhibitors also to compounds that modulate RAS-MAPK signalling, in rare as well as in common, non-syndromic forms of ASDs, especially in the presence of increased brain size.

Our study has some limitations. We did not have access to a cohort where we could assess the enrichment of mTOR-related *de novo* variant jointly in macrocephaly and ASDs. Therefore, these analyses were performed in two separate, phenotypically distinct cohorts. We used the largest most powerful meta-analyses for ASD genetics, that include all genome-wide genetic datasets of ASDs. Therefore, there is no availability of an independent replication dataset. The consistency between different datasets, however all point to a link between mTOR-related genes and ASDs and/or brain size. We are limited by the current knowledge and understanding of mTOR signalling and its contributing genes for gene-set and gene-set stratified analyses. Lastly, our results cannot claim causality, as to translate genetic association findings to more mechanistic molecular understanding, it would require the use of *in vitro* or *in vivo* models. Such experiments would help to establish if variation in this mTOR gene-set indeed represent a causal factor for large brains in ASDs.

Our findings demonstrate that a set of mTOR-related genes associate with both ASDs and brain volume, and that variation in mTOR underlies a positive genetic correlation between them. Our findings suggest that the mTOR pathway is a biological mediator of the phenotypic association of ASDs and large brains.

## Data Availability

all data produced in the present work are available upon reasonable request to the authors

## Supplementary tables (excel spreadsheet)

Table S1. List of genes included in the full mTOR gene-set and its sub-branches

Table S2. *De novo* variants in genes of the mTOR gene-set in the ID cohort

Table S3. *De novo* variants in mTOR-related genes in individuals with autism from the *de-novo-db*

Table S4. Sensitivity analyses of *de novo* variants in individuals with autism for the two mTOR sub-branches and restricted to brain-expressed genes in the *de-novo-db*

## Acknowledgements

This project has received funding from the European Union’s Horizon 2020 research and innovation programme under grant agreement No. 847879 (PRIME, Prevention and Remediation of Insulin Multimorbidity in Europe). This publication is part of the project ‘No labels needed: understanding psychiatric disorders based on genetic traits’ (with project number 09150161910091) of the research program Veni, which is partly financed by the Dutch Research Council (NWO) Health Research and Development (ZonMW). The analyses were carried out on the Dutch national e-infrastructure, and it is part of the research programme Computing Time National Computing Facilities Processing Round pilots 2018 with project No. 17666, which is (partly) financed by the Dutch Research Council (NWO).

Brunner H.G. is in part supported by the Solve-RD project that has received funding from European Community’s Horizon 2020 research and innovation programme under agreement No. 779257. Mota R.N. is supported by European Community’s Horizon 2020 programme (H2020/2014-2020) under agreement No. 667302 (CoCA) and by funding from the Dutch National Science Agenda NeurolabNL project (grant 400-17-602).

## Ethical considerations

For the local ID cohort, diagnostic WES was approved by the medical ethics committees of Radboudumc (Commissie Mensegebonden Onderzoek; registration number 2011-188). Written informed consent was obtained from all the families. Other datasets used in the current manuscript are based on publicly available datasets. In de-novo db all participants in each study provided written informed consents and agreed with the use of their data for research purposes. All the individuals included in the PGC, iPSYCH, ENIGMA and CHARGE cohort samples provided written informed consent.

#### Key points

- Autism spectrum disorders (ASDs) and brain volume show a phenotypic association, but no genetic correlation has been reported
- We show that rare and common variants in genes from the mTOR signalling pathway associate to both ASDs and brain volume
- Using a stratified genetic correlation approach, restricting to variants within mTOR-related genes, we show a positive genetic correlation between ASDs and brain volume
- By dissecting the genetic relationship by function we can delineate potential biological mechanisms between complex traits, like ASDs and brain volume.

## Supporting information

### ASD IPSYCH -PGC GWAS sample

Summary statistics of the ASD GWAS came from the meta-analysis of the iPSYCH ASD sample and the PGC ASD samples. These samples consist of individuals of Caucasian, European ancestry. Summary statistics from the meta-analysis can be retrieved through the PGC website https://www.med.unc.edu/pgc/about-us/.

The iPSYCH ASD sample is a population-based case-control sample extracted from a baseline cohort consisting of all children born in Denmark between 1981 and 2005. Cases were identified from the Danish Psychiatric Central Research Register (DPCRR), which includes data on all individuals treated in Denmark at psychiatric hospitals. ASD diagnosis was based on ICD10 criteria, and include diagnosis for childhood autism, atypical autism, Asperger’s syndrome, other pervasive neurodevelopmental disorders and/or unspecified.

Children without an ASD diagnosis were defined as controls. The PGC ASD samples includes 5 ASD case-control samples: The Geschwind Autism Center of Excellence (ACE; n = 391), the Autism Genome Project (AGP; n = 2272), the Autism Genetic Resource Exchange (AGRE; n = 974), the Montreal/Boston Collection (MONBOS; n = 1396), and the Simons Simplex Collection (SSC; n = 2231). For further details, see the PGC website: https://www.med.unc.edu/pgc/files/resultfiles/PGCASDEuro_Mar2015.readme.pdf. Here, only the CEU subset was selected based on the results of principal component analyses. All participants provided written informed consent.

### ICV ENIGMA-CHARGE cohort

ICV GWAS data were based on the analysis of 25,974 individuals from 46 independent studies belonging to the Enhancing Neuroimaging Genetics Through Meta-analysis (ENIGMA) consortium (N= 13,171) and Cohorts for Heart and Aging Research in Genomic Epidemiology

(CHARGE) consortium (N = 12,803). The summary statistics of the meta-analysis of these two projects can be retrieved through the ENIGMA website http://enigma.ini.usc.edu/research/download-enigma-gwas-results/

The ENIGMA consortium includes samples with neuroimaging data in a range of neurological and neuropsychiatric disorders and in control populations. The CHARGE consortium includes mainly population-based studies investigating the underpinnings of health related traits and age-related disorders. Only individuals of Caucasian, European ancestry were taken along for analyses. All participants provided written informed consent.

### ID cohort diagnostic WES analyses

Since September 2011 whole exome sequencing (WES) is part of the routine diagnostic work-up aimed at the identification of the genetic causes underlying intellectual disability (ID) diseases at the Radboud University Medical Center and Maastricht University Medical Center. For individuals with unexplained ID, a family-based WES approach is used which allows the identification of de novo variants as well as variants segregating according to other types of inheritance, including recessive variants and maternally inherited X-linked recessive variants in males. For this study, we selected all individuals with ID who had data from family-based WES in the time period of 2013-2018. This selection yielded a set of 2,418 individual probands, including 1040 females and 1378 males across 2387 different families. Of these, 161 were excluded because of lack of data, no ID or termination of pregnancy. The level of ID ranged between mild (IQ 50-70) to severe-profound (IQ<30). Families gave informed consent for both the diagnostic procedure as well as for forthcoming research that could result in the identification of new genes underlying ID by meta-analysis. The exomes of 2,257 patient-parent trios were sequenced, using DNA isolated from blood, at the Beijing Genomics Institute (BGI) in Copenhagen. Exome capture was performed using Agilent SureSelect v4 and v5 and samples were sequenced on an Illumina HiSeq 4000 instrument with paired-end reads to a median target coverage of 112x. Sequence reads were aligned to the hg19 reference genome using BWA version v0.7.12 and duplicate marking by Picard v1.90. Variants were subsequently called by the GATK haplotypecaller (version v3.4-46). The diagnostic WES process as outlined above only reports (de novo) variants that can be linked to the individuals’ phenotype. In this study, we systematically collected all de novo variants in regions in or close to (200bp) a capture target. De novo variants were called as described previously. Briefly, variants called within parental samples were removed from the variants called in the child. For the remaining variants pileups were generated from the alignments of the child and both parents. Based on pileup results variants were then classified into the following categories: “maternal (when identified in the mother only)”, “paternal (when identified in the father only)”, “low coverage” (when there was insufficient read depth in either parent), “shared” (when identified in both parents)”, and “possibly de novo” (when absent in the parents). Variants classified as possibly de novo were included in this study. We applied various quality filters to ensure that only the most reliable calls were included in the study, based on stringent quality control. For this quality control we selected on GATK Quality score > 450, minimal number of variant reads of 10, minimal number of total reads of 20, minimal percentage of variant reads of 20%, frequency in dbSNP < 0.1%, coverage in parents of at least 10 reads, and we discarded 15 complex variants after manual inspection in the Integrative Genomics Viewer (https://igv.org/app/).

## References

Adams, H. H. H., Hibar, D. P., Chouraki, V., Stein, J. L., Nyquist, P. A., Rentería, M. E., Trompet, S., Arias-Vasquez, A., Seshadri, S., Desrivières, S., Beecham, A. H., Jahanshad, N., Wittfeld, K., van der Lee, S. J., Abramovic, L., Alhusaini, S., Amin, N., Andersson, M., Arfanakis, K., … Thompson, P. M. (2016). Novel genetic loci underlying human intracranial volume identified through genome-wide association. Nature Neuroscience, 19(12), 1569–1582. https://doi.org/10.1038/nn.4398

American Psychiatric Association. (2013). Diagnostic and Statistical Manual of Mental Disorders, 5.

Brien, H. E. O., Hannon, E., Hill, M. J., Toste, C. C., Robertson, M. J., Morgan, J. E., Mclaughlin, G., Lewis, C. M., Schalkwyk, L. C., Hall, L. S., Pardiñas, A. F., Owen, M. J., Donovan, M. C. O., Mill, J., & Bray, N. J. (2018). Expression quantitative trait loci in the developing human brain and their enrichment in neuropsychiatric disorders. Genome Biology, 19(194), 1–13.

Bulik-Sullivan, B., Loh, P. R., Finucane, H. K., Ripke, S., Yang, J., Patterson, N., Daly, M. J., Price, A. L., Neale, B. M., Corvin, A., Walters, J. T. R., Farh, K. H., Holmans, P. A., Lee, P., Collier, D. A., Huang, H., Pers, T. H., Agartz, I., Agerbo, E., … O’Donovan, M. C. (2015). LD score regression distinguishes confounding from polygenicity in genome-wide association studies. Nature Genetics, 47(3), 291–295. https://doi.org/10.1038/ng.3211

Butler, M. G., Dazouki, M. J., Zhou, X. P., Talebizadeh, Z., Brown, M., Takahashi, T. N., Miles, J. H., Wang, C. H., Stratton, R., Pilarski, R., & Eng, C. (2005). Subset of individuals with autism spectrum disorders and extreme macrocephaly associated with germline PTEN tumour suppressor gene mutations. Journal of Medical Genetics, 42(4), 318–321. https://doi.org/10.1136/jmg.2004.024646

Costa-Mattioli, M., & Monteggia, L. M. (2013). mTOR complexes in neurodevelopmental and neuropsychiatric disorders. Nature Neuroscience, 16(11), 1537–1543. https://doi.org/10.1038/nn.3546

de Leeuw, C. A., Mooij, J. M., Heskes, T., & Posthuma, D. (2015). MAGMA: Generalized Gene-Set Analysis of GWAS Data. PLoS Computational Biology, 11(4), 1–19. https://doi.org/10.1371/journal.pcbi.1004219

Fidler, D. J., Bailey, J. N., & Smalley, S. L. (2000). Macrocephaly in autism and other pervasive developmental disorders. Developmental Medicine and Child Neurology, 42(11), 737–740. https://doi.org/10.1017/S0012162200001365

Ganesan, H., Balasubramanian, V., Iyer, M., Venugopal, A., Subramaniam, M. D., Cho, S. G., & Vellingiri, B. (2019). mTOR signalling pathway - A root cause for idiopathic autism? BMB Reports, 52(7), 424–433. https://doi.org/10.5483/BMBRep.2019.52.7.137

Gerring, Z. F., Mina-Vargas, A., Gamazon, E. R., & Derks, E. M. (2021). E-MAGMA: an eQTL-informed method to identify risk genes using genome-wide association study summary statistics. Bioinformatics, 37(16), 2245–2249. https://doi.org/10.1093/bioinformatics/btab115

Grasby, K. L., Jahanshad, N., Painter, J. N., Colodro-Conde, L., Bralten, J., Hibar, D. P., Lind, P. A., Pizzagalli, F., Ching, C. R. K., McMahon, M. A. B., Shatokhina, N., Zsembik, L. C. P., Thomopoulos, S. I., Zhu, A. H., Strike, L. T., Agartz, I., Alhusaini, S., Almeida, M. A. A., Alnæs, D., … Medland, S. E. (2020). The genetic architecture of the human cerebral cortex. Science, 367(6484). https://doi.org/10.1126/science.aay6690

Grove, J., Ripke, S., Als, T. D., Mattheisen, M., Walters, R. K., Won, H., Pallesen, J., Agerbo, E., Andreassen, O. A., Anney, R., Awashti, S., Belliveau, R., Bettella, F., Buxbaum, J. D., Bybjerg-Grauholm, J., Bækvad-Hansen, M., Cerrato, F., Chambert, K., Christensen, J. H., … Børglum, A. D. (2019). Identification of common genetic risk variants for autism spectrum disorder. Nature Genetics, 51(3), 431–444. https://doi.org/10.1038/s41588-019-0344-8

Huber, K. M., Klann, E., Costa-Mattioli, M., & Zukin, R. S. (2015). Dysregulation of Mammalian Target of Rapamycin Signaling in Mouse Models of Autism. Journal of Neuroscience, 35(41), 13836–13842. https://doi.org/10.1523/jneurosci.2656-15.2015

Kanehisa, M., & Goto, S. (2000). KEGG: Kyoto Encyclopedia of Genes and Genomes. Nucleic Acids Research, 28, 27–30. https://doi.org/10.3892/ol.2020.11439

Kim, W. (2015). Brain size is controlled by the mammalian target ofrapamycin (mTOR) in mice. Communicative & Integrative Biology, February, 11–13.

Klein, S., Sharifi-Hannauer, P., & Martinez-Agosto, J. A. (2013). Macrocephaly as a clinical indicator of genetic subtypes in autism. Autism Research, 6(1), 51–56. https://doi.org/10.1002/aur.1266

Kumar, V., Zhang, M. X., Swank, M. W., Kunz, J., & Wu, G. Y. (2005). Regulation of dendritic morphogenesis by Ras-PI3K-Akt-mTOR and Ras-MAPK signaling pathways. Journal of Neuroscience, 25(49), 11288–11299. https://doi.org/10.1523/JNEUROSCI.2284-05.2005

Laplante, M., & Sabatini, D. (2012). mTOR signaling in growth control and disease Mathieu. Cell, 149(2), 274–293. https://doi.org/10.1016/j.cell.2012.03.017.mTOR

Licausi, F., & Hartman, N. W. (2018). Role of mTOR complexes in neurogenesis. International Journal of Molecular Sciences, 19(5). https://doi.org/10.3390/ijms19051544

Lindley, A. A., Benson, J. E., Grimes, C., Cole, T. M., & Herman, A. A. (1999). The relationship in neonates between clinically measured head circumference and brain volume estimated from head CT-scans. Early Human Development, 56(1), 17–29. https://doi.org/10.1016/S0378-3782(99)00033-X

Lu, Q., Li, B., Ou, D., Erlendsdottir, M., Powles, R. L., Jiang, T., Hu, Y., Chang, D., Jin, C., Dai, W., He, Q., Liu, Z., Mukherjee, S., Crane, P. K., & Zhao, H. (2017). A Powerful Approach to Estimating Annotation-Stratified Genetic Covariance via GWAS Summary Statistics. American Journal of Human Genetics, 101(6), 939–964. https://doi.org/10.1016/j.ajhg.2017.11.001

Onore, C., Yang, H., Van de Water, J., & Ashwood, P. (2017). Dynamic Akt/mTOR Signaling in Children with Autism Spectrum Disorder. Frontiers in Pediatrics, 5(March), 1–9. https://doi.org/10.3389/fped.2017.00043

Reijnders, M. R. F., Kousi, M., van Woerden, G. M., Klein, M., Bralten, J., Mancini, G. M. S., van Essen, T., Proietti-Onori, M., Smeets, E. E. J., van Gastel, M., Stegmann, A. P. A., Stevens, S. J. C., Lelieveld, S. H., Gilissen, C., Pfundt, R., Tan, P. L., Kleefstra, T., Franke, B., Elgersma, Y., … Brunner, H. G. (2017). Variation in a range of mTOR-related genes associates with intracranial volume and intellectual disability. Nature Communications, 8(1), 1–11. https://doi.org/10.1038/s41467-017-00933-6

Sacco, R., Gabriele, S., & Persico, A. M. (2015). Head circumference and brain size in autism spectrum disorder: A systematic review and meta-analysis. Psychiatry Research - Neuroimaging, 234(2), 239–251. https://doi.org/10.1016/j.pscychresns.2015.08.016

Schneider, M., de Vries, P. J., Schönig, K., Rößner, V., & Waltereit, R. (2017). mTOR inhibitor reverses autistic-like social deficit behaviours in adult rats with both Tsc2 haploinsufficiency and developmental status epilepticus. European Archives of Psychiatry and Clinical Neuroscience, 267(5), 455–463. https://doi.org/10.1007/s00406-016-0703-8

Shimobayashi, M., & Hall, M. N. (2014). Making new contacts: The mTOR network in metabolism and signalling crosstalk. Nature Reviews Molecular Cell Biology, 15(3), 155–162. https://doi.org/10.1038/nrm3757

Sun, T., & Hevner, R. (2014). Growth and folding of the mammalian cerebral cortex: from molecules to malformations. Nat Rev Neuroscience, 15(4), 217–232. https://doi.org/10.1038/nrn3707

The Autism Spectrum Disorders Working Group of The Psychiatric Genomics Consortium. (2017). Meta-analysis of GWAS of over 16,000 individuals with autism spectrum disorder highlights a novel locus at 10q24.32 and a significant overlap with schizophrenia. Molecular Autism, 8, 21. https://doi.org/10.1186/s13229-017-0137-9

Tick, B., Bolton, P., Happé, F., Rutter, M., & Rijsdijk, F. (2016). Heritability of autism spectrum disorders: A meta-analysis of twin studies. Journal of Child Psychology and Psychiatry and Allied Disciplines, 57(5), 585–595. https://doi.org/10.1111/jcpp.12499

Tilot, A. K., Bebek, G., Niazi, F., Altemus, J. B., Romigh, T., Frazier, T. W., & Eng, C. (2016). Neural transcriptome of constitutional Pten dysfunction in mice and its relevance to human idiopathic autism spectrum disorder. Molecular Psychiatry, 21(1), 118–125. https://doi.org/10.1038/mp.2015.17

Turner, T. N., Yi, Q., Krumm, N., Huddleston, J., Hoekzema, K., Stessman, H. A. F., Doebley, A. L., Bernier, R. A., Nickerson, D. A., & Eichler, E. E. (2017). denovo-db: A compendium of human de novo variants. Nucleic Acids Research, 45(D1), D804–D811. https://doi.org/10.1093/nar/gkw865

Van Rooij, D., Anagnostou, E., Arango, C., Auzias, G., Behrmann, M., Busatto, G. F., Calderoni, S., Daly, E., Deruelle, C., Di Martino, A., Dinstein, I., Duran, F. L. S., Durston, S., Ecker, C., Fair, D., Fedor, J., Fitzgerald, J., Freitag, C. M., Gallagher, L., … Buitelaar, J. K. (2018). Cortical and subcortical brain morphometry differences between patients with autism spectrum disorder and healthy individuals across the lifespan: Results from the ENIGMA ASD working group. American Journal of Psychiatry, 175(4), 359–369. https://doi.org/10.1176/appi.ajp.2017.17010100

Vonder Haar, C., Martens, K. M., Riparip, L. K., Rosi, S., Wellington, C. L., & Winstanley, C. A. (2017). Frontal Traumatic Brain Injury Increases Impulsive Decision Making in Rats: A Potential Role for the Inflammatory Cytokine Interleukin-12. Journal of Neurotrauma, 34(19), 2790–2800. https://doi.org/10.1089/neu.2016.4813

Wen, Y., Alshikho, M. J., & Herbert, M. R. (2016). Pathway network analyses for autism reveal multisystem involvement, major overlaps with other diseases and convergence upon MAPK and calcium signaling. PLoS ONE, 11(4), 1–23. https://doi.org/10.1371/journal.pone.0153329

Werme, J., van der Sluis, S., Posthuma, D., & de Leeuw, C. A. (2021). LAVA: An integrated framework for local genetic correlation analysis. BioRxiv, 2020.12.31.424652. https://doi.org/10.1101/2020.12.31.424652

Yeung, K. S., Tso, W. W. Y., Ip, J. J. K., Mak, C. C. Y., Leung, G. K. C., Tsang, M. H. Y., Ying, D., Pei, S. L. C., Lee, S. L., Yang, W., & Chung, B. H. Y. (2017). Identification of mutations in the PI3K-AKT-mTOR signalling pathway in patients with macrocephaly and developmental delay and/or autism. Molecular Autism, 8(1), 1–11. https://doi.org/10.1186/s13229-017-0182-4

Yu, J. S. L., & Cui, W. (2016). Proliferation, survival and metabolism: The role of PI3K/AKT/ mTOR signalling in pluripotency and cell fate determination. Development (Cambridge), 143(17), 3050–3060. https://doi.org/10.1242/dev.137075

Zhao, B., Ibrahim, J. G., Li, Y., Li, T., Wang, Y., Shan, Y., Zhu, Z., Zhou, F., Zhang, J., Huang, C., Liao, H., Yang, L., Thompson, P. M., & Zhu, H. (2019). Heritability of Regional Brain Volumes in Large-Scale Neuroimaging and Genetic Studies. Cerebral Cortex, 29(7), 2904–2914. https://doi.org/10.1093/cercor/bhy157

